# Symptom persistence and biomarkers in post-COVID-19/chronic fatigue syndrome – results from a prospective observational cohort

**DOI:** 10.1101/2023.04.15.23288582

**Authors:** A. F. Legler, L. Meyer-Arndt, L. Mödl, C. Kedor, H. Freitag, E. Stein, U. Hoppmann, R. Rust, Frank Konietschke, A. Thiel, F. Paul, C. Scheibenbogen, J. Bellmann-Strobl

**Affiliations:** Charité – Universitätsmedizin Berlin, corporate member of Freie Universität Berlin, Humboldt Universität zu Berlin and Berlin Institute of Health, Max Delbrück for Molecular Medicine, Experimental and Clinical Research Center, 13125 Berlin, Germany; Charité – Universitätsmedizin Berlin, corporate member of Freie Universität Berlin, Humboldt Universität zu Berlin and Berlin Institute of Health, NeuroCure Research Center, 10117 Berlin, Germany; Charité – Universitätsmedizin Berlin, corporate member of Freie Universität Berlin, Humboldt Universität zu Berlin and Berlin Institute of Health, Department for Neurology with Experimental Neurology, 10117 Berlin, Germany; Charité - Universitätsmedizin Berlin, corporate member of Freie Universität Berlin and Humboldt-Universität zu Berlin and Institute of Biometry and Clinical Epidemiology, 10117 Berlin, Germany; Charité – Universitätsmedizin Berlin, corporate member of Freie Universität Berlin, Humboldt Universität zu Berlin and Berlin Institute of Health, Institute of Medical Immunology, 13353 Berlin, Germany; Charité - Universitätsmedizin Berlin, corporate member of Freie Universität Berlin, Humboldt-Universität zu Berlin, and Berlin Institute of Health, Regenerative Immunology and Aging, BIH Center for Regenerative Therapies, 13353 Berlin, Germany; Si-M / “Der Simulierte Mensch” a science framework of Technische Universität Berlin and Charité - Universitätsmedizin Berlin, 10117 Berlin, Germany

## Abstract

**Introduction:** Post-COVID-19 syndrome (PCS) is characterized by a wide range of symptoms, predominantly fatigue and exertional intolerance. While disease courses during the first year post infection have been repeatedly described, little is known about long-term health consequences.

**Methods:** We assessed symptom severity and various biomarkers at three time points post infection (3-8 months (mo), 9-16mo, 17-20mo) in 106 PCS patients with moderate to severe fatigue and exertional intolerance. A subset of patients fulfilled diagnostic criteria of myalgic encephalomyelitis/chronic fatigue syndrome (PCS-ME/CFS) based on the Canadian Consensus Criteria.

**Results:** While PCS-ME/CFS patients showed persisting symptom severity and disability up to 20mo post infection, PCS patients reported an overall health improvement. Inflammatory biomarkers equally decreased in both groups. Lower hand grip force at onset correlated with symptom persistence especially in PCS-ME/CFS.

**Discussion:** Debilitating PCS may persist beyond 20mo post infection, particularly in patients fulfilling diagnostic criteria for ME/CFS.

## Introduction

Post-COVID-19 syndrome (PCS) is worldwide recognized as sequela of coronavirus disease 19 (COVID-19) caused by severe acute respiratory syndrome coronavirus type 2 (SARS-CoV-2)^1^. There is a worrying number of patients with various persistent symptoms following mild or moderate COVID-19 mainly presenting as fatigue, exertion intolerance, headache, myalgia, neurological and cognitive deficits as well as orthostatic disturbances, which can severely impact the patients’ quality of life.^2–7^ Reports estimate a proportion of 2%-10% of all COVID-19 patients to be impaired one year after infection.^2,3,8^

More than two years into the SARS-CoV-2 pandemic, the WHO led the way to standardize the definition of PCS as part of the WHO International Classification of Diseases (ICD-10)^4^: The post-COVID condition occurs within three months after a probable SARS-CoV-2 Infection, lasts for at least two months with an impact on everyday functioning and cannot be explained by alternative diagnoses. Acknowledging the post-COVID condition was a crucial first step towards recognizing and improving the health care situation of the patients affected. While the short- and medium-term clinical presentations of PCS between 3 and 9 months after SARS-CoV-2 infection have been concisely described,^3,5,9^ little is known to date about potential long-term health consequences that may prevail beyond 12 months.

We reported previously on the first results of our ongoing prospective observational cohort study initiated in August 2020 in order to characterize patients with persisting debilitating fatigue and exertion intolerance following COVID-19.^9^ Our first analyses revealed that a subset of PCS patients developed the full scope of myalgic encephalomyelitis/chronic fatigue syndrome (ME/CFS) 6 months after initial infection^9^. ME/CFS is a complex multisystemic disease with an estimated pre-pandemic worldwide prevalence of 0.2 to 0.8%. Approximately 3 million people were diagnosed with ME/CFS in Europe alone by 2020.^10,11^ It is characterized by pronounced fatigue and post exertional malaise (PEM), cognitive impairment, orthostatic intolerance, pain and sleep disturbance while lacking evidence of macroscopic organ damage. The key symptom of ME/CFS is an intolerance to mental and physical exertion, which triggers PEM.^12^ Infections with various pathogens may cause ME/CFS such as Epstein-Barr virus (EBV), enteroviruses, human herpes virus (HHV)-6, dengue viruses, intracellular bacteria and SARS-CoV-1.^13^ The pathomechanism is still only partially elucidated: Infection-triggered autoimmunity, viral mimicry, latent virus reactivation, and autonomous dysfunction including a dysregulation in ß2-adrenergic vasoconstriction are concepts currently debated.^14^ Specifically for PCS-ME/CFS, studies have provided first evidence that autoantibodies to G protein–coupled receptors and endothelial dysfunction may play a role.^15^ Of interest, reactivation of EBV during COVID-19 frequently occurs and is considered a risk factor for developing PCS. As infectious mononucleosis caused by late primary EBV infection is a well-established trigger of ME/CFS, it is tempting to speculate that in a subset of PCS EBV may trigger the disease. ^13,16–19^Here we present the follow-up data up to 20 months after SARS-CoV-2 infection and identify predictive factors for the disease course. We hypothesized that the subgroup of PCS-ME/CFS patients develops a chronic condition with distinct clinical and paraclinical features.

## Results

### Cohort and symptom characteristics

We examined a total of 106 patients suffering from persistent moderate to severe fatigue and exertion intolerance 6 months post COVID-19 at up to two follow-up time points (9-16 months, and 17-20 months) after SARS-CoV-2 infection **(Supp. 1 Fig. 1**). 55 patients fulfilled the Canadian Consensus Criteria (CCC) for ME/CFS and are referred to as PCS-ME/CFS; the remaining 51 patients are referred to as PCS. For demographic characteristics see **Table 1**. Symptoms frequently reported by both post-COVID-19 and ME/CFS patients are shown in **Table 2** assessing symptom prevalence, severity, and evolution over time for both cohorts in detail.

**Table 1:** Demographic participant characteristics. Median and range are reported for both cohorts separately as well as all participants together.

|  | PCS (n=51) |  | PCS-ME/CFS (n=55) |  | Total (n=106) |  |
| --- | --- | --- | --- | --- | --- | --- |
|  | Median | Range | Median | Range | Median | Range |
| Sex (m/f) | 15/36 |  | 6/49 |  | 21/85 |  |
| Age | 43 | 19-66 | 43 | 20-62 | 40 | 19-66 |
| BMI | 25 | 18-40 | 24 | 15-35 | 24 | 15-40 |

**Table 2:** Frequency (in %) and severity (median scores) of symptoms as quantified by the Canadian Consensus Criteria (qCCC) for all three time periods. Group differences are reported as p-values and significant results (< 0.05) are marked in bold.

| Canadian Consensus<br>Criteria quantified (CCCq) | Baseline |  |  |  |  | Follow-up 1 |  |  |  |  | Follow-up 2 |  |  |  |  | PCS time<br>effect* | PCS-CFS/ME<br>time effect* |
| --- | --- | --- | --- | --- | --- | --- | --- | --- | --- | --- | --- | --- | --- | --- | --- | --- | --- |
|  | PCS |  | PCS-ME/CFS |  | P-value<br>between<br>groups | PCS |  | PCS-ME/CFS |  | P-value<br>between<br>groups | PCS |  | PCS-ME/CFS |  | P-value<br>between<br>groups |  |  |
|  | Median | % | Median | % |  | Median | % | Median | % |  | Median | % | Median | % |  |  |  |
| Fatigue | 7 | 100 | 8 | 100 | 0.18 | 6 | 98 | 8 | 100 | < 0.01 | 5 | 100 | 8 | 100 | < 0.01 | <0.01 | 0.58 |
| PEM | 7 | 88 | 8 | 100 | < 0.01 | 6 | 98 | 8 | 100 | < 0.01 | 5 | 96 | 8 | 98 | < 0.01 | <0.01 | 0.04 |
| Need for rest | 8 | 96 | 8 | 100 | 0.13 | 7 | 87 | 8 | 100 | < 0.01 | 6 | 100 | 8 | 100 | < 0.01 | <0.01 | 0.51 |
| Impaired performance | 8 | 98 | 8 | 100 | 0.20 | 6 | 89 | 8 | 100 | < 0.01 | 6 | 96 | 8 | 96 | 0.07 | <0.01 | 0.18 |
| Stress intolerance | 8 | 94 | 8 | 98 | < 0.01 | 7 | 97 | 8 | 100 | < 0.01 | 6 | 100 | 8 | 98 | < 0.01 | 0.04 | 0.97 |
| Muscle pain | 5 | 78 | 6 | 85 | 0.10 | 4 | 98 | 6 | 83 | 0.10 | 3 | 79 | 6 | 90 | 0.01 | 0.60 | 0.91 |
| Headache | 5 | 82 | 6 | 93 | 0.16 | 4 | 83 | 5 | 96 | 0.16 | 4 | 77.7 | 5 | 98 | 0.01 | <0.01 | 0.19 |
| Joint pain | 4 | 69 | 5 | 64 | 0.29 | 2 | 63 | 5 | 77 | 0.29 | 3 | 59 | 5 | 65 | 0.29 | 0.75 | 0.75 |
| Memory/word finding problems | 5 | 88 | 5 | 98 | 0.72 | 5 | 91 | 5 | 91 | 0.91 | 3 | 83 | 6 | 88 | 0.15 | <0.01 | 0.46 |
| Concentration impairment | 6 | 94 | 7 | 100 | 0.60 | 6 | 93 | 7 | 98 | 0.60 | 5 | 86 | 7 | 98 | < 0.01 | <0.01 | 0.62 |
| Mental Fatigue | 7 | 96 | 7 | 100 | 0.92 | 6 | 95 | 7 | 100 | 0.16 | 5 | 100 | 8 | 97 | < 0.01 | <0.01 | 0.86 |
| Visual disturbance | 3 | 66 | 3 | 67 | 1 | 2.5 | 71 | 3 | 64 | 1 | 2 | 69 | 2 | 66 | 0.90 | 0.81 | 0.81 |
| Mood change | 5 | 88.2 | 5 | 89 | 0.79 | 5 | 89 | 5 | 85 | 0.79 | 4 | 79 | 5 | 88 | 0.36 | <0.01 | 0.73 |
| Reading concentration | 5 | 90 | 6 | 100 | 0.19 | 5 | 91 | 6 | 96 | 0.19 | 3 | 86 | 6 | 90 | 0.01 | <0.01 | 0.81 |
| Palpitations | 3 | 65 | 3 | 89 | 0.08 | 2.5 | 74 | 5 | 85 | 0.07 | 3 | 76 | 5 | 80 | 0.04 | 0.49 | 0.49 |
| Standing up dizziness | 4 | 75 | 5 | 76 | 1 | 3 | 71 | 3 | 74 | 1 | 2 | 65 | 3 | 69 | 0.42 | 0.05 | 0.40 |
| Walking dizziness | 2 | 63 | 4 | 76 | 0.36 | 2 | 65 | 3 | 74 | 0.62 | 2 | 59 | 4 | 66 | 0.08 | 0.29 | 0.29 |
| Sleep disturbance | 7 | 86 | 7 | 91 | 1 | 6 | 82 | 6 | 89 | 1 | 5 | 76 | 7 | 90 | < 0.01 | <0.01 | 0.68 |
| Temperature hypersensitivity | 4 | 57 | 5 | 84 | 0.33 | 4 | 66 | 5 | 82 | 0.33 | 3 | 65 | 6 | 90 | 0.01 | 0.42 | 0.16 |
| Light hypersensitivity | 2 | 63 | 4 | 65 | 0.19 | 2 | 56 | 3 | 83 | 0.06 | 2 | 65 | 5 | 66 | 0.02 | 0.52 | 0.52 |
| Noise hypersensitivity | 5 | 73 | 6 | 91 | 0.27 | 6 | 76 | 6 | 94 | 0.41 | 4 | 96 | 7 | 90 | 0.06 | 0.43 | 0.55 |
| Breathing difficulty | 5 | 72.5 | 6 | 80 | 0.34 | 3 | 65 | 5 | 76 | 0.25 | 4 | 82 | 5 | 78 | 0.32 | 0.06 | 0.15 |
| Irritable bowel | 4 | 55 | 5 | 74 | 0.51 | 3 | 64 | 4 | 80 | 0.51 | 2 | 51.8 | 4 | 66 | 0.01 | 0.02 | 0.66 |
| Fever | 1 | 20 | 1 | 16 | 0.76 | 1 | 17 | 1 | 25 | 0.66 | 1 | 10 | 1 | 23 | 0.47 | 0.50 | 0.50 |
| Painful lymph nodes | 1 | 30 | 1 | 27 | 1 | 1 | 20 | 1 | 28 | 1 | 1 | 14 | 1 | 42 | 0.04 | 0.18 | 0.18 |
| Sore throat | 2 | 55 | 2 | 62 | 0.50 | 1 | 39 | 3 | 62 | 0.13 | 1 | 31 | 3 | 68 | < 0.01 | 0.01 | 0.29 |
| Flu-like symptoms | 4 | 74.5 | 6 | 83 | 0.04 | 2 | 52 | 5 | 71 | < 0.01 | 1.5 | 50 | 5 | 85 | < 0.01 | <0.01 | 0.30 |
| Symptom severity | 7 | 88.2 | 8 | 91 | 0.02 | 6 | 97 | 7 | 100 | 0.01 | 5 | 100 | 7 | 100 | < 0.01 | <0.01 | 0.29 |

### Fatigue and PEM remain key symptoms in PCS-ME/CFS after 18 months

Fatigue and PEM are among the key symptoms of PCS and indispensable for the diagnoses of ME/CFS. Patients diagnosed with PCS-ME/CFS were significantly more affected by fatigue than PCS patients over the entire study period. At baseline, PCS patients presented on average lower Chalder Fatigue Scale (CFQ) scores than PCS-ME/CFS patients with 33 % of PCS and 55% of PCS-ME/CFS reporting severe (≥ 28 points) fatigue (**Fig. 1a and Supp. 1 Table 1**). While CFQ scores remained on a similarly high level in the PCS-ME/CFS cohort, they significantly decreased in PCS patients. At follow-up 2, only 3% of PCS but 46 % of PCS-ME/CFS patients still reported CFQ scores ≥ 28 points. The course of symptom severity was similar for the Chalder sub-score of physical fatigue (**Fig. 1b**), the Short Form (36) (SF-36) Health Questionnaire fatigue score (**Fig. 1c)** and the fatigue score of the quantitative CCC (qCCC; **Supp. 1 Fig. 2a**). Mental fatigue slightly improved also in PCS-ME/CFS over time (**Fig. 1d**).

**Fig. 1:**
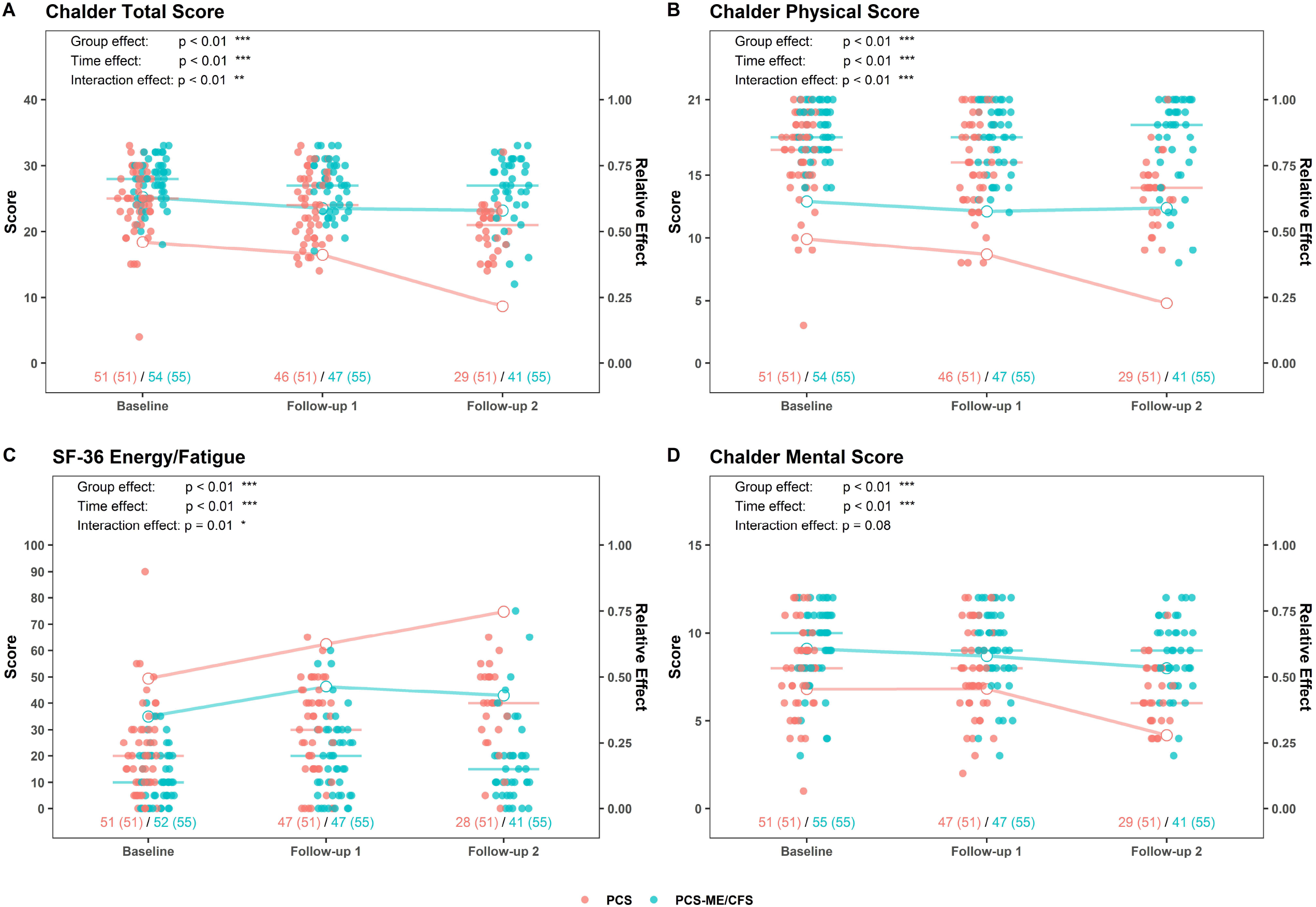
Fatigue. **a**, Chalder Fatigue Total Score, ranging from 0 (no fatigue) to 33 (severe fatigue); **b**, Chalder Fatigue Physical Score (range 0-21); **c**, SF-36 Energy/Fatigue, ranging from 0 (most impaired) to 100 (no impairment); **d**, Chalder Fatigue Mental Score (range 0-12). Dots represent absolute score values (red for PCS, blue for PCS-ME/CFS) as quantified on the left Y axis. Bars depict group medians. Lines (red for PCS, blue for PCS-ME/CFS) depict main relative time, group, and interaction effects as quantified on the right Y axis. p ≤ 0.05 = *, p ≤ 0.01=**, p ≤ 0.001=***, p ≤ 0.0001=****.

PEM duration, severity and frequency were all significantly higher in PCS-ME/CFS compared to PCS patients at baseline and remained at a higher level up to follow-up 2 (**Fig. 2a-c**). However, neither of the two cohorts experienced a reduction of PEM duration over time but an improvement in PEM frequency and severity. Remarkably, in the PCS-ME/CFS cohort, PEM duration decreased below 10 hours in 7 individuals (17%) at follow-up 2 (thus no longer fulfilling CCC) (**Fig. 2a**). Five of these seven individuals showed an equally strong improvement in PEM severity and frequency.

**Fig. 2:**
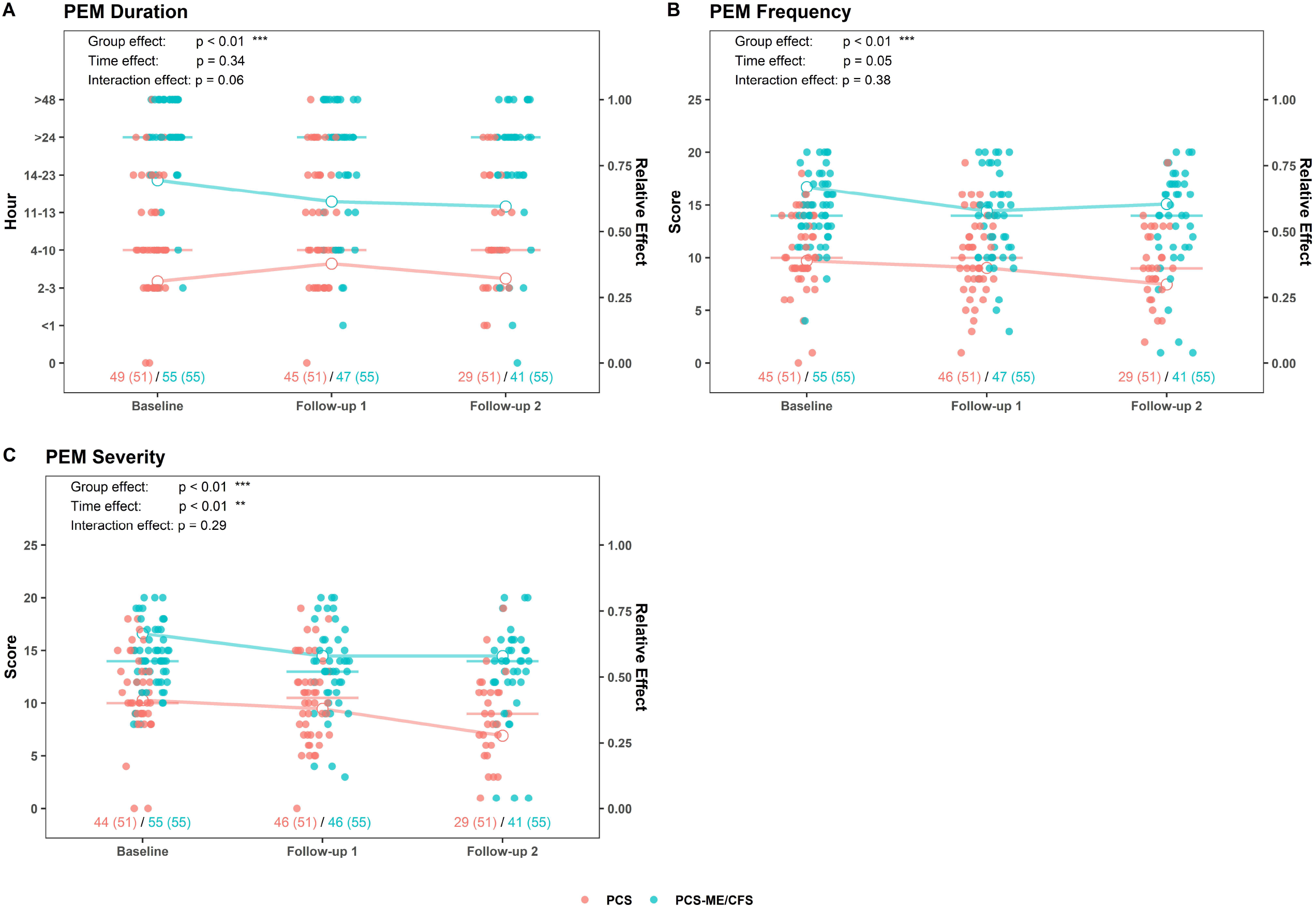
PEM. **a**, PEM duration (in hours), **b**, frequency (stated as occurring: never, rarely, half of the time, most of the time, always), and **c**, severity (stated as: not at all, mild, moderate, severe, very severe) of exhaustion experienced post exertion. Dots represent absolute score values (red for PCS, blue for PCS-ME/CFS) as quantified on the left Y axis. Bars depict group medians. Lines (red for PCS, blue for PCS-ME/CFS) depict main relative time, group and interaction effects as quantified on the right Y axis. p ≤ 0.05 = *, p ≤ 0.01=**, p ≤ 0.001=***, p ≤ 0.0001=****.

### Reduced functional disability in both cohorts over time

At baseline, both PCS and PCS-ME/CFS patients had a low median Bell disability score of 40 with 4/55 PCS-ME/CFS patients and 1/51 PCS patients reporting an inability to leave the house (Bell score 20; **Fig. 3a**). 53 of 55 PCS-ME/CFS and 43 of 51 PCS reported that they are unable to work full- or part-time (Bell score < 70/100). At follow-up 2, Bell score remained at 40 in PCS-ME/CFS but increased to 60 in PCS with only 12% of PCS-ME/CFS (5/41) but 43% of PCS patients reporting a Bell score ≥ 70. Patient reports on performance capacity as assessed with the qCCC confirmed this course (**Supp. 1 Fig. 2b**). Again, the seven PCS-ME/CFS patients who reported less PEM duration over time also considerably improved on the Bell score (**Fig. 3a**).

**Fig. 3:**
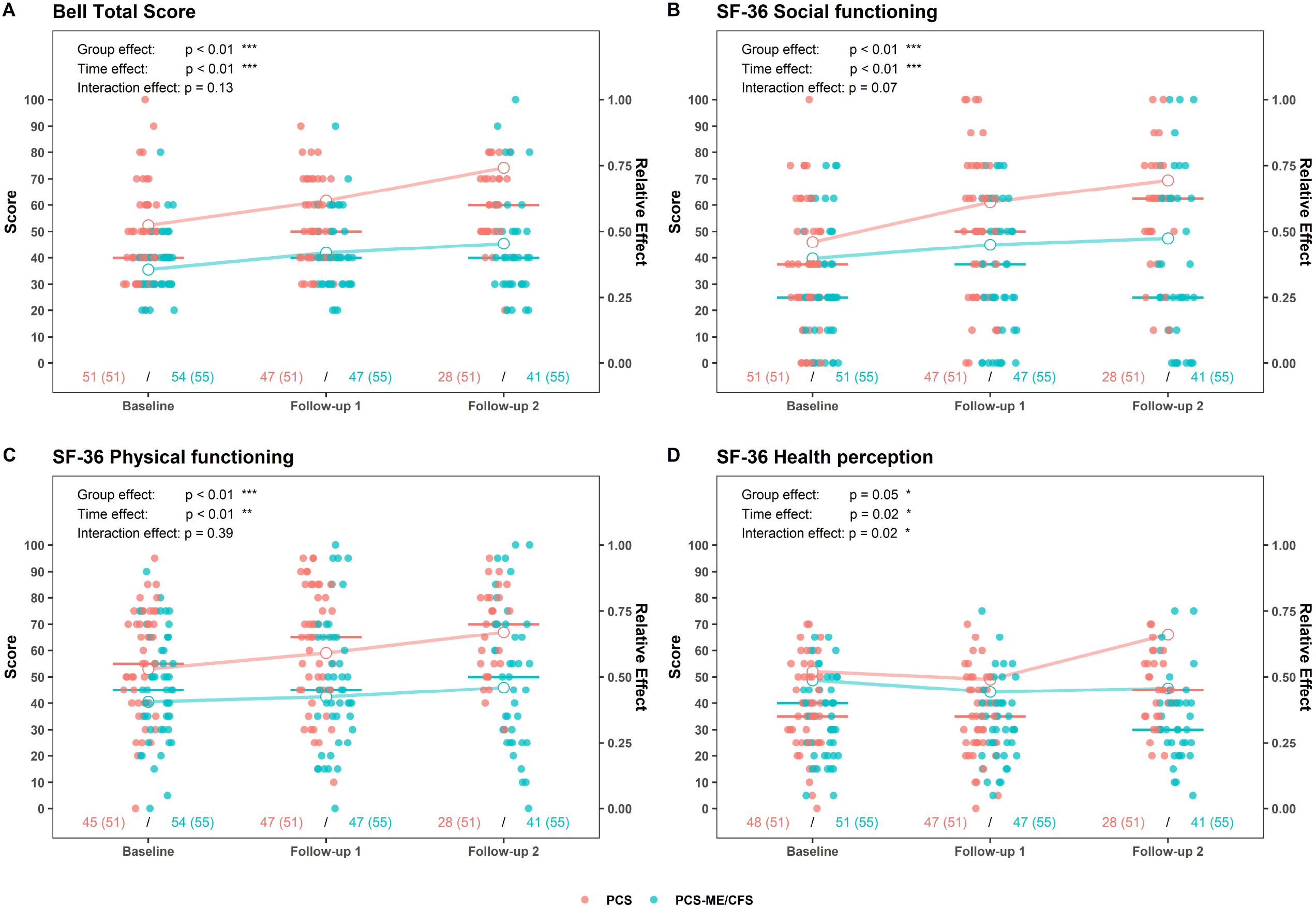
General health and functional impairment. **a**, Bell score assessing disability due to chronic fatigue from 100 (no symptoms present) to 50 (moderate symptoms at rest and moderate to severe symptoms with exercise or activity) to 0 points (unable to get out of bed independently); **b**, SF-36 social functioning; **c**, SF-36 physical functioning; **d**, SF-36 health perception, 0 points (greatest possible health limitation) - 100 points (no health limitation). Dots represent absolute score values (red for PCS, blue for PCS-ME/CFS) as quantified on the left Y axis. Bars depict group medians. Lines (red for PCS, blue for PCS-ME/CFS) depict main relative time, group, and interaction effects as quantified on the right Y axis. p ≤ 0.05 = *, p ≤ 0.01=**, p ≤ 0.001=***, p ≤ 0.0001=****.

Various sub-scores of the SF-36 were lower in PCS-ME/CFS compared to PCS at baseline (**Supp. 1 Table 1**) and group differences increased over time, mainly due to higher scores in PCS at both follow-ups (**Fig. 3a-d**). Specifically, perception of physical functioning (SF-36) was more reduced in PCS-ME/CFS compared to PCS patients from the beginning with only minor improvement at follow-up 2 in PCS-ME/CFS (median 45-50) and moderate in PCS (median 55-70) (**Fig. 3c**). Despite group differences in fatigue (**Fig. 1a-c**), PEM (**Fig. 2a-c**) and (perception of) functional disability (**Fig. 3a, c**), overall health perception as measured by the SF-36 was equally reduced to levels below 50% in both cohorts at baseline (**Fig. 3d**). Taken together, while PCS reported improved health perception up to follow-up 2, PCS-ME/CFS patients stagnated at their initial level.

### Emotional well-being improves in PCS patients only

Perception of emotional well-being as measured by the SF-36 was equally reduced by 50% in both cohorts at baseline (**Fig. 4a**). Further, no differences in the Patient Health Questionnaire 9 (PHQ9), which is used as a screening tool for affective disorders, was found at baseline (**Fig. 4b**). Seven patients (6 PCS-ME/CFS; 1 PCS) reached more than 20 points on the PHQ9 score at baseline, which suggests a PCS-associated affective burden. However, the PHQ9 includes symptoms of fatigue, cognition and sleep and thus has a low specificity for depression in PCS and ME/CFS. Impairment according to the PHQ9 improved in PCS patients from baseline to follow-up 2 but remained largely unchanged in the PCS-ME/CFS cohort (**Fig. 4a and b**). Together, these scores indicate a considerable emotional burden due to illness, which over time improves in PCS patients while PCS-ME/CFS patients remain severely impaired.

**Fig. 4:**
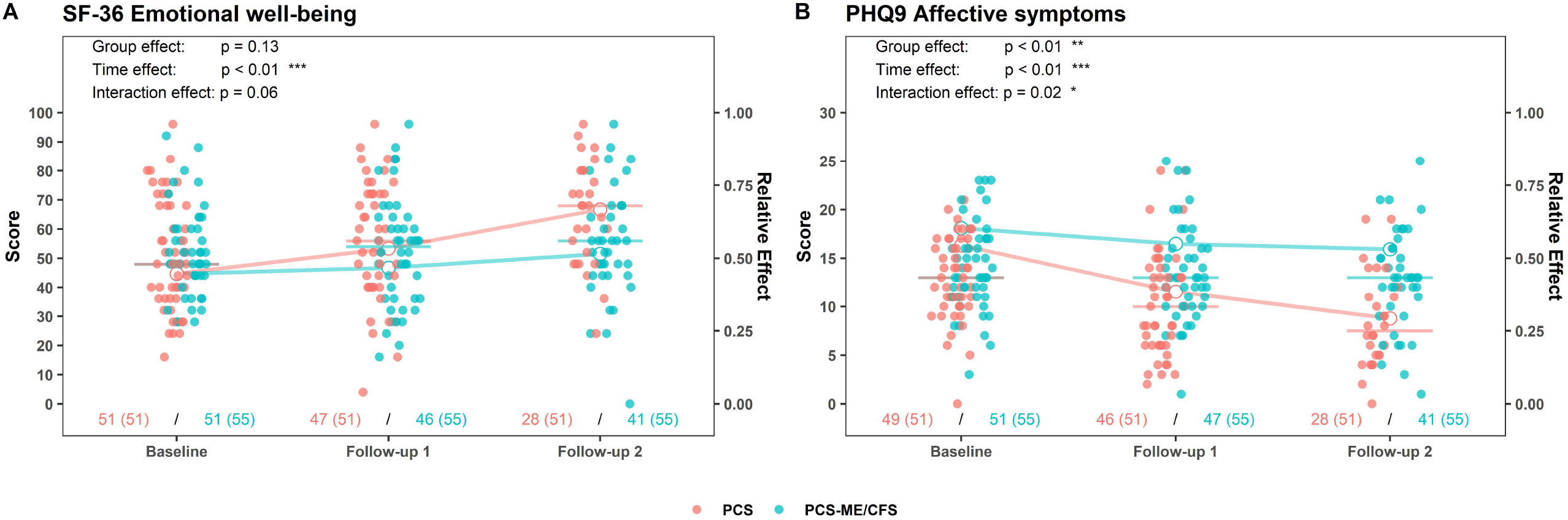
Emotional impairment. **a**, SF-36 emotional well-being, 0 points (greatest possible health limitation) - 100 points (no health limitation); **b**, Patient Health Questionnaire 9 (PHQ9), minimal depressive symptoms (1-4), mild depressive symptoms (5-9), moderate depressive symptoms (10-14), moderately severe depressive symptoms (15-19), or severe depressive symptoms (20-27). Dots represent absolute score values (red for PCS, blue for PCS-ME/CFS) as quantified on the left Y axis. Bars depict group medians. Lines (red for PCS, blue for PCS-ME/CFS) depict main relative time, group, and interaction effects as quantified on the right Y axis. p ≤ 0.05 = *, p ≤ 0.01=**, p ≤ 0.001=***, p ≤ 0.0001=****.

### PCS-ME/CFS remain more severely affected by pain than PCS

According to the SF-36 pain score, PCS-ME/CFS patients were more affected by pain than PCS patients at all time points and improved in the PCS cohort from baseline to follow-up 2 while there was just a minor improvement among PCS-ME/CFS patients (**Fig. 5a**). In detail, muscle pain (qCCC) was reported by 85% of PCS-ME/CFS patients and 78% of PCS patients at baseline while joint pain (qCCC) was reported by 64% of PCS-ME/CFS patients and 69% of PCS patients (**Table 2, Supp. 1 Fig. 2c and d**). None of these symptoms improved significantly over time in either of the two cohorts: Both patient groups reported joint and muscle pain with a severity of more than 5/10 up to follow-up 2. Headaches were mentioned by 82% PCS and 93% PCS-ME/CFS patients at baseline with a decreasing severity over time in both cohorts (**Supp. 1 Fig. 2e**).

**Fig. 5:**
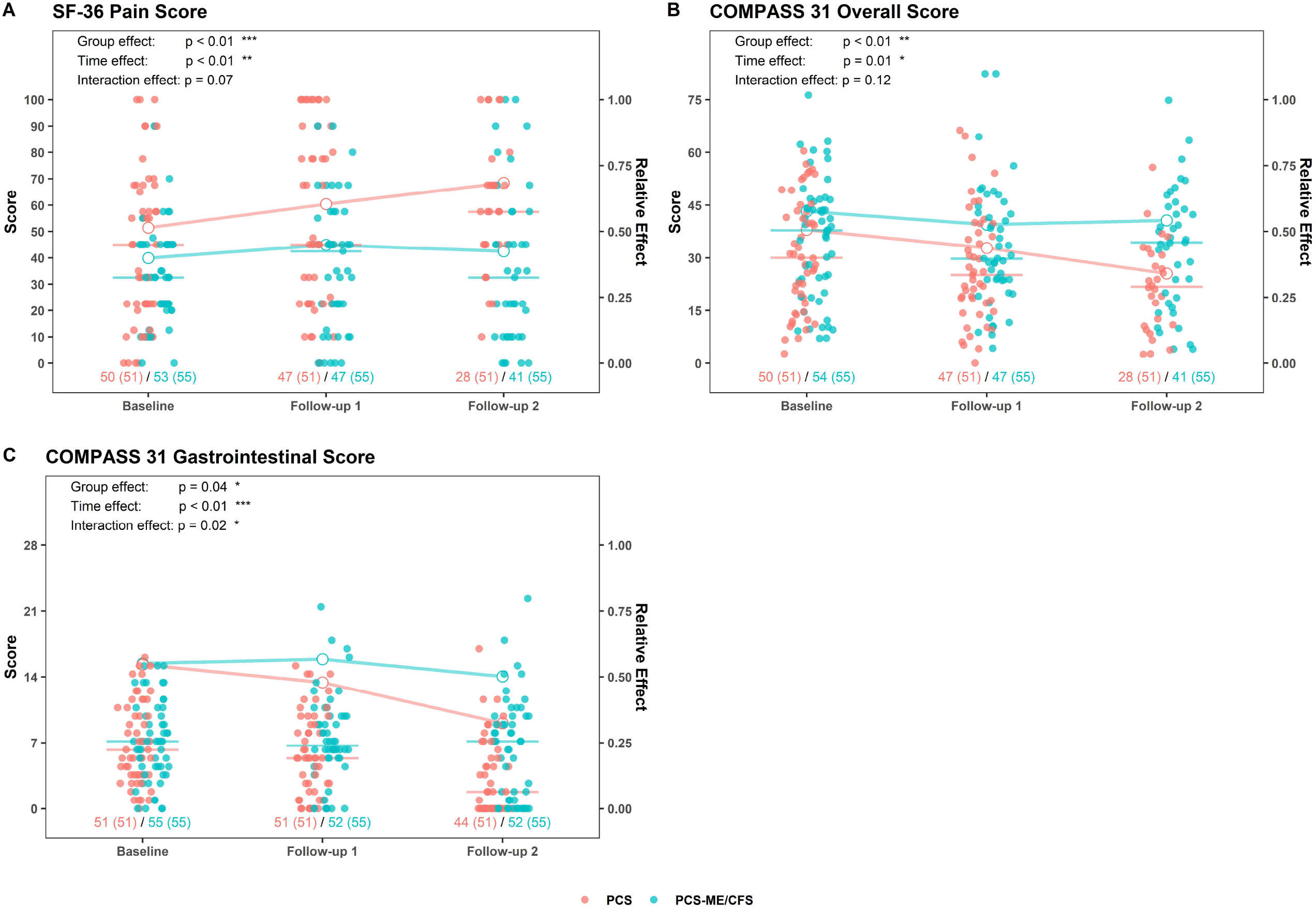
Pain and autonomic dysfunction. **a**, SF-36 pain score, 0 points (greatest possible health limitation) - 100 points (no health limitation); **b**, COMPASS 31 overall score, 0 (no symptoms) - 100 (severe autonomic dysfunction); **c**, COMPASS 31 gastrointestinal score (range 0-25). Dots represent absolute score values (red for PCS, blue for PCS-ME/CFS) as quantified on the left Y axis. Bars depict group medians. Lines (red for PCS, blue for PCS-ME/CFS) depict main relative time, group, and interaction effects as quantified on the right Y axis. p ≤ 0.05 = *, p ≤ 0.01=**, p ≤ 0.001=***, p ≤ 0.0001=****.

### Prevalence of neurological symptoms remains at a high level in both cohorts

At baseline, the prevalence of cognitive symptoms such as concentration (**Supp. 1 Fig. 2f**) and memory/wordfinding difficulties (**Table 2**), mental fatigue (**Supp. 1 Fig. 2g**), and difficulties while reading (**Table 2**) was comparable in both cohorts. Assessing symptom evolution over time, overall cognitive impairment as summarized in the qCCC (**Supp. 1 Fig. 2h**) ameliorated solely in PCS patients. Hypersensitivities to noise, light and temperature, which are characteristic symptoms in ME/CFS were more frequent in PC-ME/CFS (**Table 2**) and all of them were more pronounced in PCS-ME/CFS at second follow-up. High median symptom severity scores at follow-up 2 indicated a continuous burden from neurological impairment despite overall improvement (**Table 2**).

### Clinical signs of ongoing inflammation persist in PCS-ME/CFS patients

Patients suffering from postinfectious fatigue syndromes often state persisting flu-like symptoms, painful lymph nodes and a sore throat, which can be signs of ongoing inflammation. These symptoms were here summed up as qCCC immune score (**Supp. 1 Fig. 2i**), which was equally elevated in both cohorts at baseline. However, symptoms significantly decreased in PCS patients but persisted in PCS-ME/CFS patients (**Supp. 1 Fig. 2i**).

### Regression of autonomic dysfunctions in both PCS and PCS-ME/CFS patients

Autonomic dysfunction is a common feature of postinfectious fatigue syndromes. At baseline, PCS and PCS-ME/CFS patients showed comparable signs of autonomic dysfunction reflected by a Composite Autonomic Symptom Score 31 (COMPASS 31) overall score indicative of moderate to severe complaints. Despite an improvement of this overall score over time in both cohorts, PCS-ME/CFS patients were more affected at second follow-up than PCS (**Fig. 5b and Supp. 1 Tab. 1**). Gastrointestinal complains only improved in PCS patients with an almost unchanged level of impairment in PCS-ME/CFS at follow-up (**Fig. 5c and Supp. 1 Fig. 2j**). On a similar note, PCS-ME/CFS patients continued to suffer from more severe sleep disturbances compared to PCS (qCCC; **Supp. 1 Fig. 2k**).

To further characterize autonomic dysfunction in our patients and investigate potential clinical implications, we measured the adaption of blood pressure and pulse to postural change. At baseline, 7% (5/42) of PCS and 11% (5/44) of PCS-ME/CFS patients showed signs of postural tachycardia syndrome (POTS) and 4% (2/42) of PCS and 6% (3/44) of PCS-ME/CFS showed signs of orthostatic hypotension (OH). Up to follow-up 2, none of the PCS patients continued to show signs of POTS or OH (but two patients showed newly emerged symptoms of OH or POTS at follow-up 1). In contrary, 6% (3/44) of PCS-ME/CFS presented POTS symptoms also at follow-up 2 suggesting a persistent autonomic dysfunction in this cohort (one patient showed new signs of OH).

### Persistent diminished HGS but improvement of muscle fatiguability over time

Hand grip strength (HGS), a reliable parameter to quantify frailty and mortality, was evaluated in in two consecutive sessions at baseline (for female patients only to avoid gender bias and due to an insufficient number of male patients, n=35 PCS, n=49 PCS-ME/CFS) and follow-up 1 (n=25 PCS, n=37 PCS-ME/CFS; **Fig. 6**) and results were compared to age-dependent reference values as reported by Jäkel et al.^29^ We found no significant differences in mean (fmean) and maximum force (fmax) between PCS and PCS-ME/CFS and no changes in HGS over time in either cohort. A remarkable number of PCS (63%) and PCS-ME/CFS patients (67%) showed measurements below their respective cut-offs, i.e., for mean force in the second session (fmean2) at baseline and follow-up 1 (PCS 40%; PCS/ME/FCS 67 %; **Fig. 6a and b**).

**Fig. 6:**
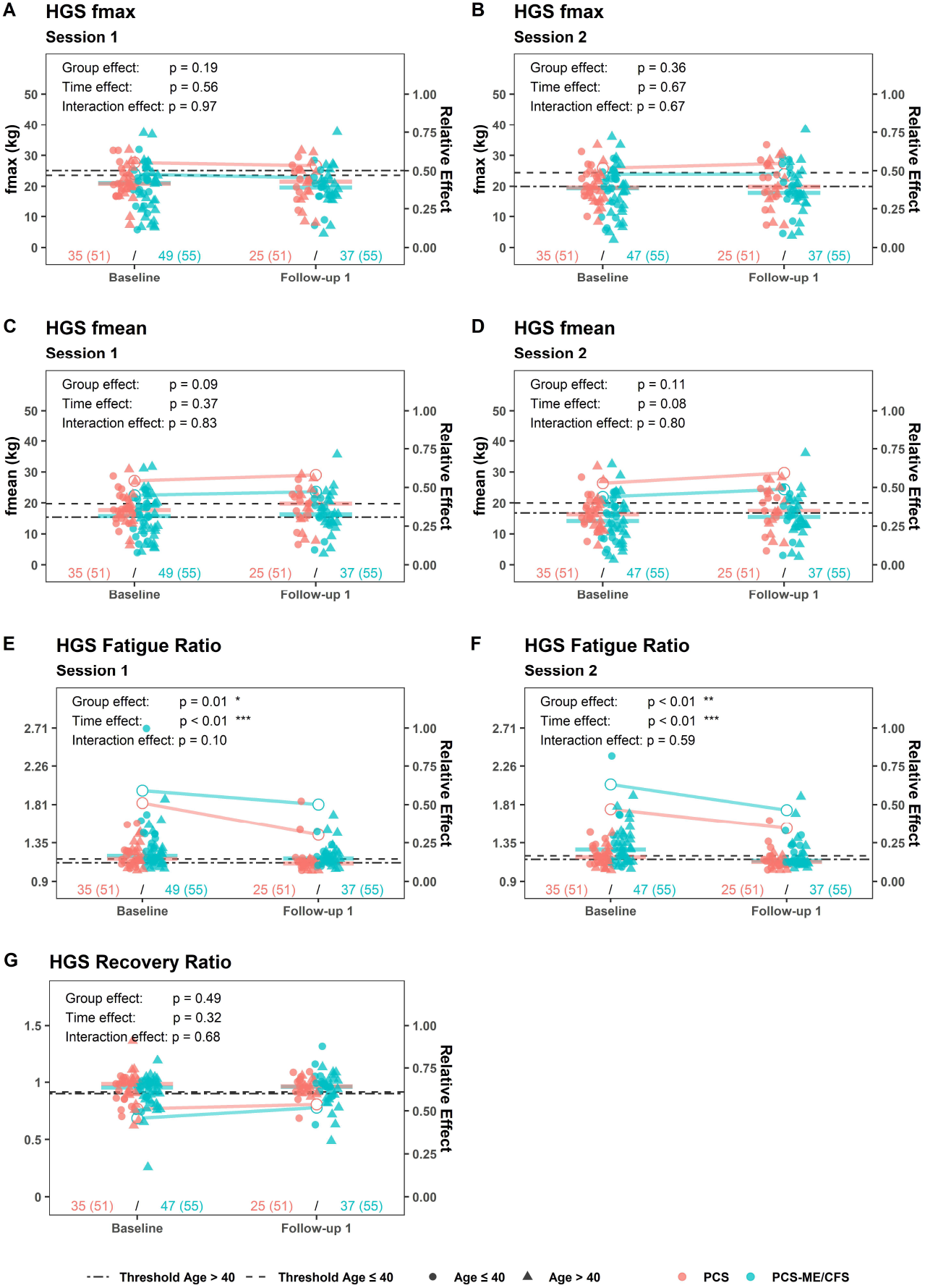
Hand grip strength. **a-d**, mean (**a and b**) and maximum (**c and d**) force in kg of all 10 measurements per session for session 1 (**a and c**) and 2 (**b and d**) for baseline and follow-up 1; **e and f**, fatigue ratio (fmax/fmean) per session for session 1 (**e**) and 2 (**f**) and for baseline and follow-up 1; **g**, recovery ratio (fmean2/fmean1) for baseline and follow-up 1. Triangle data points depict patients <40 years. Age-dependent cut-offs are depicted according to Jäkel et al.^29^ Red dots represent PCS cohort, white dots PCS-ME/CFS cohort. Bars depict group medians with a 95% confidence interval. p ≤ 0.05 = *, p ≤ 0.01=**, p ≤ 0.001=***, p ≤ 0.0001=****. Abbreviations: BL = baseline, FU1 = follow-up 1, se_1 = session 1, se_2 = session 2.

The fatigue ratio (fmax/fmean) was determined for each session as a correlate for muscle fatigability with higher values indicating a stronger decline in force (**Fig. 6e and f**).^29^ PCS-ME/CFS patients showed higher fatigue ratios at baseline (session 2) and at follow-up (session 1) than PCS patients. The fatigue ratio decreased over time in both cohorts. No changes over time (or group differences) were found for the recovery ratio, which serves as marker for muscle strength recovery after an hour (**Fig. 6g**).^29^

### Reduction of inflammation markers over time in both cohorts

We investigated a range of biomarkers linked to postinfectious fatigue in a subset of patients (PCS n=35; PCS-ME/CFS n=31). Previous studies have found that the proinflammatory and neutrophil-recruiting interleukin 8 (IL-8, CXCL8), antinuclear antibodies (ANA) and ferritin were associated with the post-COVID condition.^20–22^ Among these patients, IL-8, which was determined after erythrocyte lysis to reflect its level of production for a duration of up to 3 months, was equally elevated in almost half of both patient cohorts at baseline.^23^ Over time, IL-8 levels decreased significantly in both groups (**Fig. 7a**). Consistently, ferritin, another inflammatory marker, was equally increased in almost a third of all patients at baseline, but again decreased in both cohorts until follow-up 2 (**Fig. 7b**). Antinuclear antibodies (ANA) were detected in 25% (8/32) of PCS patients and 26% (8/30) PCS-ME/CFS patients at baseline without significant changes over time (**Fig. 7c**). Furthermore, 35% of all patients were deficient of serum phosphate (PO4) at baseline, which is associated with increased mortality in COVID-19.^24^ Up to follow-up 1, this deficiency receded in most PCS patients while persisting in PCS-ME/CFS (**Fig. 7e**).

**Fig. 7:**
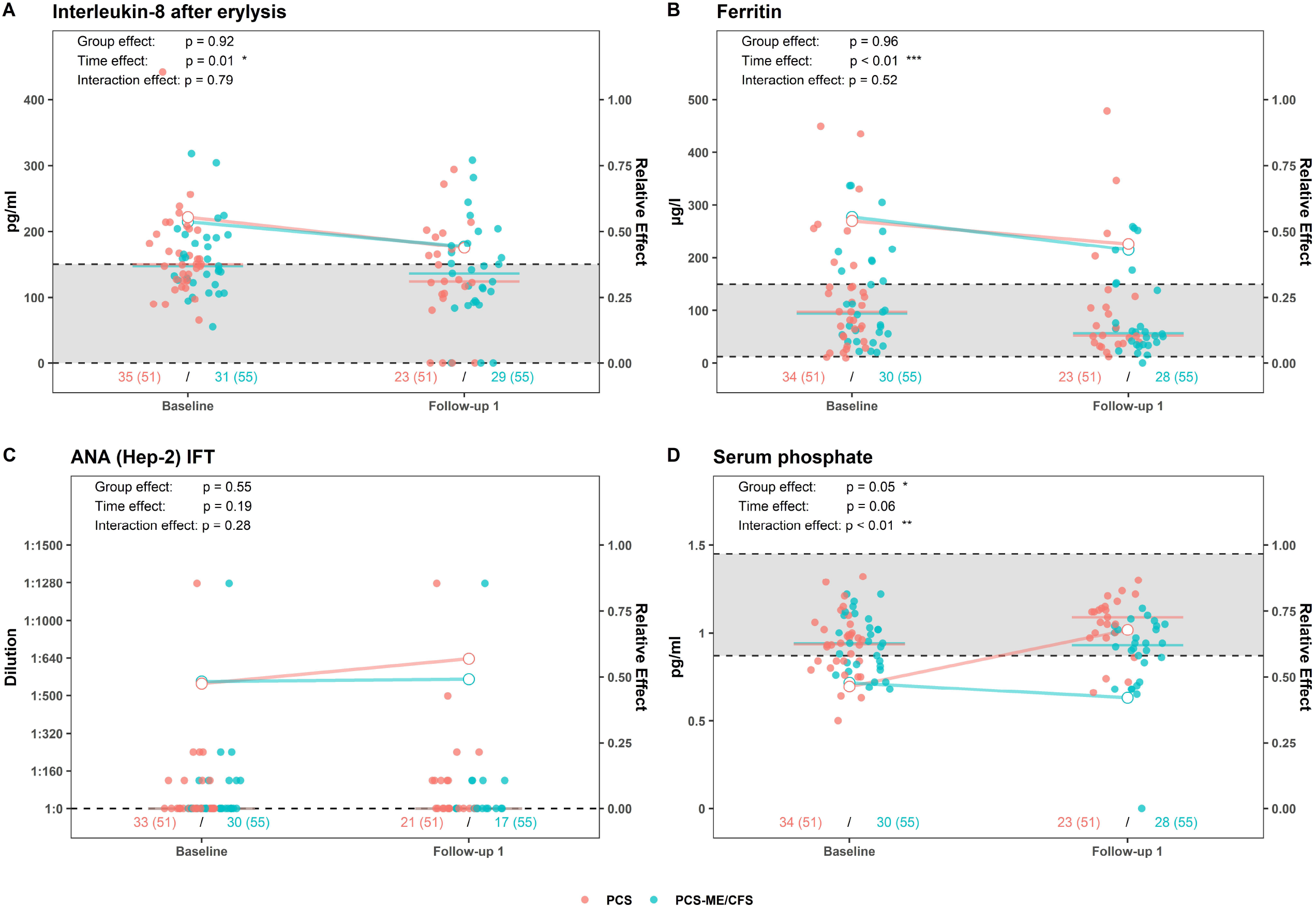
Inflammatory biomarkers associated with postinfectious fatigue. **a**) interleukin 8 (IL-8) after erylysis, reference <150 pg/ml, **b**, ferritin, reference range 13-150 μg/l, **c**, antinuclear antibodies (ANA), negative reference 1:0 dilution, **d**, serum phosphate (PO4), reference range 0.87-1.45 mmol/l, for baseline and follow-up 1 respectively. Dots represent absolute score values (red for PCS, blue for PCS-ME/CFS) as quantified on the left Y axis. Bars depict group medians. Lines (red for PCS, blue for PCS-ME/CFS) depict main relative time, group, and interaction effects as quantified on the right Y axis. p ≤ 0.05 = *, p ≤ 0.01=**, p ≤ 0.001=***, p ≤ 0.0001=****.

### Initial hand grip strength is associated with symptom burden at follow-up in PCS-ME/CFS

As disease courses and outcomes vary between individual patients, we aimed at identifying an objective marker, which could be used to estimate disease prognosis. We found HGS to be diminished in PCS-ME/CFS and to be associated with disease severity.^25^ In line with this, we observed HGS baseline measurements at baseline to strongly correlate with symptom burden in PCS-ME/CFS patients at follow-up 1: Low HGS mean and maximum force at baseline correlated with increased fatigue (CFQ), PEM, functional disability, pain, sleep disturbance and emotional impairment at follow-up 1. Consistently, a high HGS fatigue ratio indicating faster fatiguability at baseline correlated with increased fatigue (CFQ) and functional disability at follow-up 1, and low HGS recovery ratios at baseline with increased fatigue (CFQ) and disability at follow-up 2 (**Fig. 8a**). In PCS patients, we found fewer and weaker correlations of some HGS parameters at baseline with symptom outcomes at follow-up (**Fig. 8b**).

**Fig. 8:**
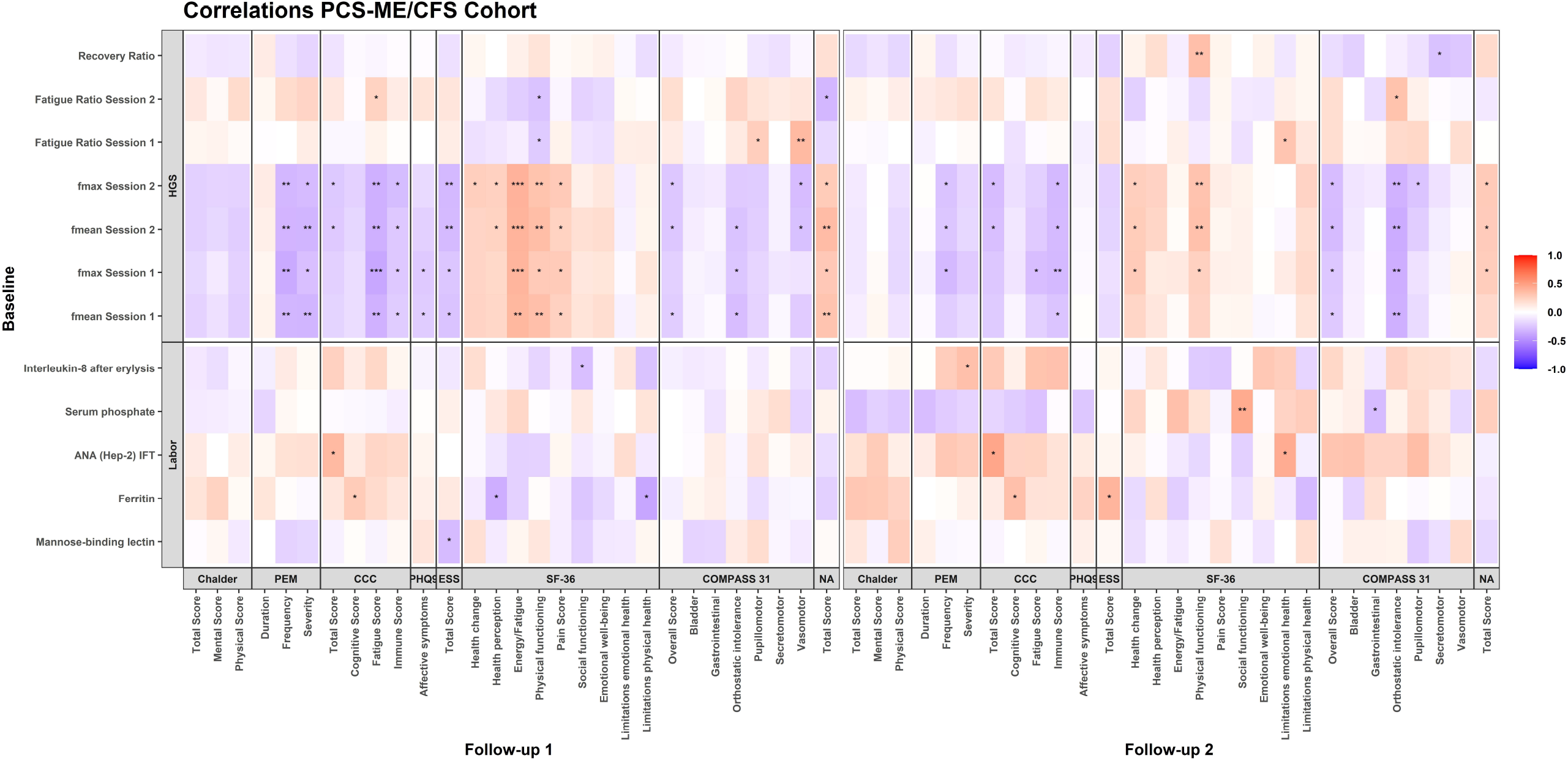
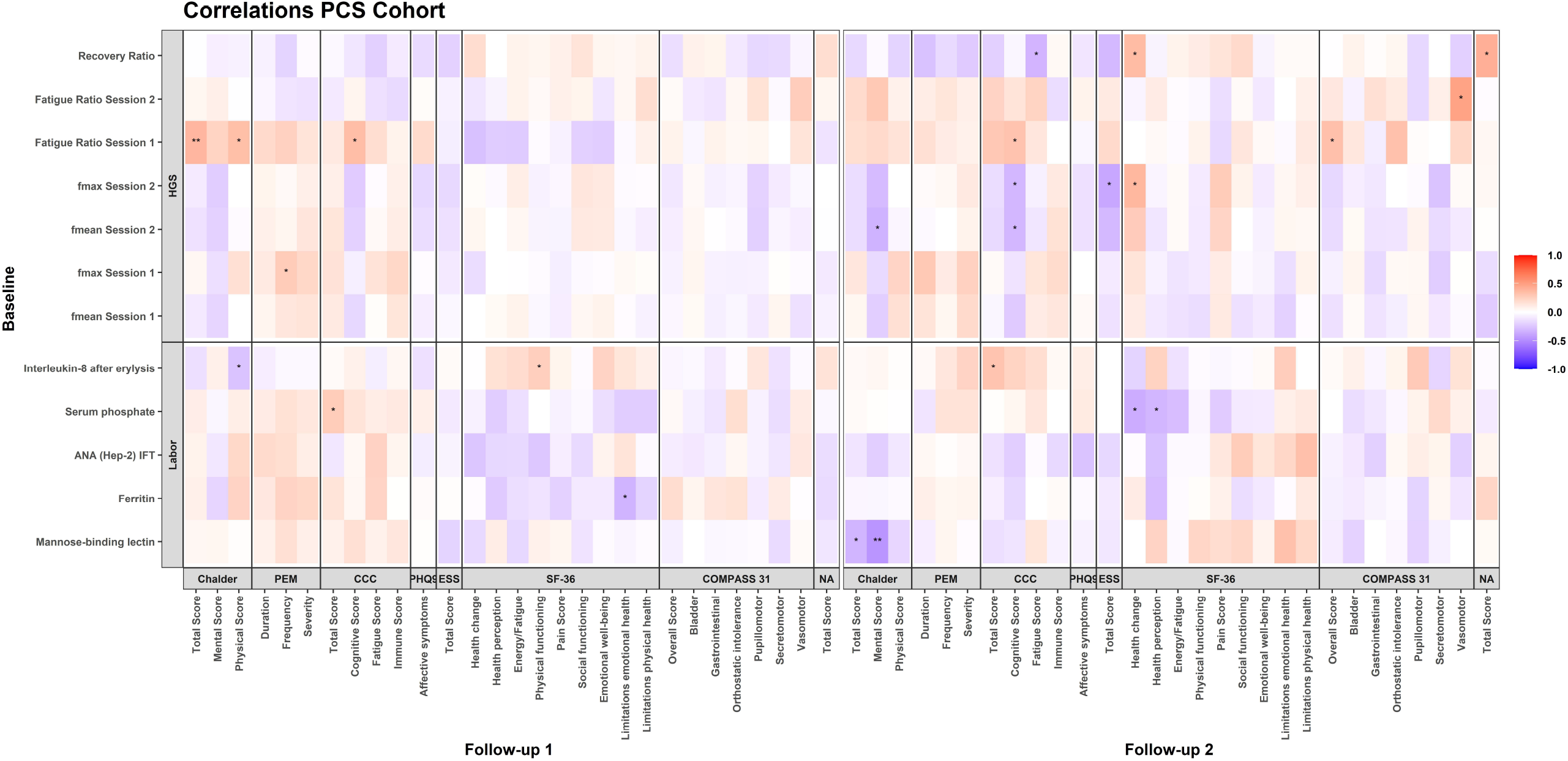
Correlations of initial symptom severity, HGS and biomarkers with longitudinal symptom severity in a, PCS-ME/CFS patients and b, PCS patients. Markers at baseline are shown on the x axis, parameters at follow-up 1 and 2 on the y axis. Blue coloring indicates negative correlations, red coloring indicates positive correlations, see color legend below the plot; p ≤ 0.05 = *, p ≤ 0.01=**, p ≤ 0.001=***, p ≤ 0.0001=****.

Next, we evaluated potential associations between biomarkers at baseline and persisting disease burden. In the PCS-ME/CFS cohort, high IL-8 levels at baseline were associated with decreased social functioning at follow-up 1 and high PEM severity at follow-up 2. High ferritin levels correlated with increased cognitive impairment at both follow-ups as well as with poor health perception and more limitation due to physical health at follow-up 1. Further, elevated ANA levels correlated with high symptom severity at follow-up 1 and 2 and low serum PO4 levels were linked to reduced social functioning and increased gastrointestinal complaints at follow-up 2. Surprisingly, in PCS patients, high IL-8 levels at baseline correlated with high total symptom scores (qCCC) at follow-up 2 but improved physical functioning and less physical fatigue (CFQ) at follow-up 1. Likewise, reduced serum PO4 was associated with low total symptom scores (qCCC) at follow-up 1 while at follow-up 2 (in line with PCS-ME/CFS) the link was to decreased health perception and health change. High MBL levels were associated with less fatigue symptoms (CFQ) at follow-up 2.

## Discussion

We here provide a comprehensive longitudinal characterization of the post-COVID-19 condition in patients with pronounced fatigue and exertion intolerance over a period of 20 months following COVID-19. All patients suffered from PCS, with a subgroup fulfilling the CCC for ME/CFS.

While both PCS and PCS-ME/CFS patients continued to report post-COVID-19 symptoms throughout the observation period^26,27^, clinical improvement was observed to variable degrees and mostly restricted to the non-ME/CFS sub-cohort. This is in line with Tran et al. (37) who monitored symptom evolution in patients with persisting symptoms after acute COVID-19 based on an online survey over a 12-month period. The proportion of patients with persisting symptoms in their cohort was about 85%. After an initial decrease, symptoms plateaued 6 to 8 months after onset.^8^ Consistently, Seeßle et al. (38) reported that neurocognitive deficits following COVID-19 can persist beyond 12 months and lead to a marked reduction of quality of life. ^28^

We here showed that patients fulfilling criteria for ME/CFS continued to be more affected than PCS patients by a wide range of symptoms including fatigue, physical disability, impaired social functioning, and emotional well-being. Importantly, exertion intolerance and PEM as the hallmark of post-infectious fatigue syndromes remained more pronounced in PCS-ME/CFS up to 20 months after initial infection. However, the extent of PEM did not improve in either cohort. Considering this persistence of PEM in most PCS patients, our study provides evidence that in ME/CFS, despite early diagnosis, prognosis is poor for most patients. Accordingly, its management by determining individual activity limits and balancing rest and activity (i.e., pacing) gains in importance. All patients in this study were seen in our specialist outpatient clinics and received recommendations for symptomatic treatment and self-management strategies. However, symptomatic therapy in ME/CFS requires prompt clinical follow-ups, which have not been available for most patients due to a lack of knowledge among most primary physicians.^29^ We are currently evaluating in a clinical trial if comprehensive care and close monitoring can improve physical function and well-being in ME/CFS patients.

The degree of fatigue and functional disability as evaluated by Chalder and Bell scales and hand grip strength and fatiguability were found to discriminate best between PCS and PCS-ME/CFS patients 16 to 20 months post infection. POTS was found only in PCS-ME/CFS patients at a prevalence of 6% consistent with other reports. Given these characteristic features and distinct disease course of the PCS-ME/CFS sub-group, classification of patients with the post-COVID condition based on the CCC is useful for further diagnostics and treatment. 16 of the 51 PCS patients not fulfilling the CCC would have fulfilled the IOM (Institute of Medicine) criteria for ME/CFS as these criteria do not predefine the length of PEM and require only fatigue, sleep disturbance and cognitive or orthostatic symptoms as mandatory symptoms. In line with other studies, patients fulfilling the CCC were more impaired and more symptomatic.^30^

The majority of patients reported newly emerged affective symptoms and poor emotional well-being after COVID-19 diagnosis, which were thus directly related to their post-COVID-19 condition. These symptoms improved only in PCS patients along with their overall clinical condition and therefore must rather be considered a consequence of the burdening disease impacting PCS-ME/CFS patients’ quality of life. Consequently, psychological support should be integrated into PCS management.

We found baseline HGS to be linked to persisting disease severity, particularly in the PCS-ME/CFS cohort. PCS-ME/CFS patients with initially reduced HGS were more likely to experience high disease burden up to 20 months after infection. Specifically, higher hand grip force correlated with a lower amount of fatigue, exertion intolerance, physical functioning and disability, the characteristic hallmarks of chronic postinfectious fatigue syndromes. In PCS patients, links of HGS to these symptom measures were not found or were much less pronounced than in PCS-ME/CFS. We thus assume that HGS is a more valuable prognostic parameter for patients with PCS-ME/CFS. Consequently, HGS measurements could serve as an easy to perform method to estimate prognosis of PCS-ME/CFS patients. However, these correlations should be considered observational as we could not control for potential confounding variables due to the limited number of participants. Therefore, these results need to be validated in further studies.

We observed a distinct improvement of symptoms (fatigue, PEM, and disability) in 7 out of 55 PCS-ME/CFS patients. These patients initially presented with severe symptoms and fulfilled the Canadian Consensus diagnostic criteria for ME/CFS. At follow-up 2, their Bell scores were improved above a value of 60 points. 5 of them also demonstrated improved hand grip strength over time. We were unable to identify common characteristics that could explain this improvement in this sub-group.

Our evaluation of biomarkers associated with post-infectious fatigue supports the presumption of ongoing inflammation in post COVID-19. The pro-inflammatory, neutrophil-recruiting cytokine IL-8 and ferritin were elevated in a subgroup of patients 6 months after infection. IL-8 levels decreased towards 12 months after SARS-CoV-2 infection. Further, we observed elevated ANA titers in 25% of patients. This is consistent with other studies that found elevated ANA titers at 12 months post-COVID, which correlated with persisting symptoms and inflammation in PCS patients. This indicates that ANA could be a relevant marker for autoimmunity in PCS.^31^ Hypophosphatemia was found in one third of patients at baseline and interestingly persisted only in the ME/CFS cohort. The etiology of hypophosphatemia is complex and potential causes are mitochondrial dysfunction with depletion of adenosine triphosphate (ATP), insulin resistance, and respiratory alkalosis.^24^ Complications of hypophosphatemia include impaired cellular ATP metabolism and increased affinity of hemoglobin to oxygen in red blood cells, which may exacerbate fatigue as well as neurologic, cardiovascular and muscle dysfunction. Thus, the effect of phosphate supplementation should be evaluated in ME/CFS. In acute COVID, results on supplementation approaches are still inconsistent.^32^

A limitation of this study is the participant drop-out during the study period. Approximately 78% of dropouts were diagnosed with PCS at baseline, the remaining dropouts with PCS-ME/CFS. As PCS patients were overall more likely to experience clinical improvement, they may have regarded further participation in the study as unbeneficial. Hence, their clinical improvement during follow-up could not be documented, which may bias the PCS cohort towards the more affected individuals. However, a great strength of this study is the comprehensive disease evaluation including an extensive and interdisciplinary set-up of questionnaires and on-site clinical examinations and functional and laboratory tests. Furthermore, more clinically less affected PCS patients would have only emphasized the discriminatory effect observed between our two patient groups.

Taken together, the post-COVID-19 condition may develop into a chronic syndrome with long-lasting symptoms and impairment. In patients fulfilling the CCC, the chance of relevant improvement by symptomatic therapy is low. Against the backdrop of over 675 million documented SARS-CoV-2 infections worldwide (status February 2023), these results suggest that the post-COVID-19 syndrome continues to present a heavy burden for those affected as well as on healthcare systems. Further studies on the pathomechanism and therapy approaches are urgently needed.

## Methods

### Study design and cohort characteristics

This work’s data was collected as part of the Pa-COVID study of the Charité – Universitätsmedizin Berlin and approved by the ethics committee of the Charité in accordance with the 1964 Declaration of Helsinki and its later amendments (EA2/006/20). The current manuscript analyses follow-up data from a prospective observational cohort study of patients with severe fatigue and exertion intolerance post COVID-19 diagnosis.^33^ Patients were recruited via the Charité’s Fatigue outpatient clinic’s website and the post-COVID consultation of the outpatient clinic of Charité’s Experimental and Clinical Resarch Center. From a total of 250 patients who were seen between July 2020 and February 2022, 171 fulfilled the inclusion criteria of **(Supp. Fig. 1a**): (1) confirmed previous diagnosis of mild to moderate COVID-19 according to WHO criteria I or II, (2) persistent moderate to severe fatigue according to the CFQ and PEM, (3) absence of COVID-19-related organ dysfunction in case of indicative symptoms and 4) absence of preexisting fatigue or relevant cardiac, respiratory, neurological, or psychiatric comorbidities according to the European Network on ME/CFS (EUROMENE) guidelines.^10^ COVID-19 diagnose was confirmed by SARS-CoV-2 polymerase chain reaction or anti-SARS-CoV-2 IgG serology (only prior to vaccination). To exclude COVID-19-related organ dysfunction, patients with severe headache or cognitive impairment were evaluated by a neurologist, patients with respiratory problems underwent comprehensive pulmonological examination including chest computer tomography and pulmonary function tests with diffusing capacity and patients with chest pain, postural tachycardia, palpitations, or elevated NT-pro BNP had a cardiological examination and were assessed by electrocardiogram (ECG), 24h ECG and echocardiography. Baseline assessment was conducted 3 to 8 months post COVID-19 manifestation. Follow-up visits were scheduled 9 to 16 months (follow-up 1) and 17 to 20 months (follow-up 2) post COVID-19 diagnosis (see **Supp. Fig. 1b**). All visits included questionnaires as well as functional and laboratory tests (**see Supp. Fig. 1b**). An additional questionnaire-based assessment was conducted 9 months post COVID-19 to standardize diagnoses of patients with baseline visits at months 3 and 4 post infection as ME/CFS can only be diagnosed after symptom persistence of more than 6 months. These assessments were not used for analysis. 65 patients (n=51 PCS, n=14 PCS-ME/CFS) dropped out during the study resulting in 106 patients with complete datasets from at least two different assessments timepoints (baseline and one follow-up).

### Diagnosis and symptom assessment

PCS was diagnosed based on fatigue, PEM and, optionally, additional key symptoms following COVID-19 persisting for at least 3 months and impairing daily live according to the WHO “clinical case definition of post-COVID-19 condition”.^4^ The diagnosis of PCS-ME/CFS was based on the CCC and PEM lasting until the next day. The minimum PEM duration required for diagnosis was set at 14 hours as this cut-off value has been shown to be reliable in distinguishing ME/CFS fatigue from fatigue associated with other diseases.^34^ In contrary, patients with PCS did not fulfill all CCC criteria and most presented with PEM less than 14 hours.

Post-COVID-19 cardinal symptoms and their severity were assessed (on a scale from 1-10, no symptoms to extreme symptoms) using the qCCC (2003).^35^ Symptom complexes were summed up respectively: Fatigue, impaired performance, need for rest and post exertional malaise were summarized as qCCC fatigue score. Painful lymph nodes, sore throat and flu-like symptoms were summed up as qCCC immune score. Concentration impairment, memory/wordfinding problems and mental fatigue were summerzied as qCCC mental score. The CFQ is a broadly recognized measuring tool for the diagnosis of ME/CFS fatigue and contains 11 items on an ordinal scale of 0 to 3 with a minimal total score of 0 (no fatigue) to a maximum total score of 33 (strong fatigue).^36,37^ The two subscales are rated as follows: mental fatigue (CFQA) contains 4 items with a range of 0 to 12 and physical fatigue (CFQB) 7 items with a range of 0 to 21. Frequency, severity and duration of PEM symptoms were assessed according to Cotler et al. (range 0 to 46, no PEM to frequent, severe or long-lasting PEM).^34^ Impairment in daily life due to chronic fatigue was rated based on the Bell disability scale with increasing impairment from 100 (no symptoms present) to 50 (moderate symptoms at rest and moderate to severe symptoms with exercise or activity; unable to perform strenuous duties, but able to perform light duty or desk work 4-5 hours a day) and 0 points (unable to get out of bed independently).^38^ Physical and social dysfunction, bodily pain, emotional well-being, general health perception and health change were evaluated by the SF-36, which is used as a generic measurement tool to assess health perception, disease progression and experienced impairment through illness.^39^ Its score ranges from 0 (greatest possible health limitation) to 100 (no relevant health limitation) points. The subitems orthostatic intolerance and gastrointestinal function of the COMPASS 31 were used to detect symptoms of autonomic dysfunction often reported by PCS patients with a minimum total score of 0 (no symptoms) and a maximum total score of 100 (strong autonomic dysfunction).^40^ Furthermore, we used the PHQ9, a questionnaire to screen for depressive symptoms^26^: Due to the overlap of several PCS symptoms with depressive disorders, the PHQ9 score was used descriptively only.

### Functional tests and biomarker assessment

POTS, OH and diminished HGS were used as clinical markers to comprehensively characterize the broad variety of symptoms and their severity seen in PCS patients. POTS and OH describe different phenotypes of autonomic dysfunction and are often seen in ME/CFS patients.^41–43^ For evaluation, blood pressure and heart rate were measured in seated position, immediately after standing up and after 2, 5 and 10 minutes (standing). Orthostatic intolerance was defined as increase of more than 30 bpm or above 120 bpm within 10 minutes after standing up.^43,44^ OH was defined as decrease of systolic pressure of more than 20 mmHg or diastolic pressure of more than 10 mmHg at any measurement.^43^ HGS, which is a meaningful marker for evaluation of muscle exertion and fatigability in fatigue patients, was assessed using an electronic dynamometer.^25^ In detail, patients gripped the measuring device 10 times with maximum force with their leading hand. This procedure was repeated after 60 minutes. Maximum (fmax1, fmax2) and mean (fmean1, fmean 2) force of each session were determined in kg. Previous studies by Jäkel et al. identified reference values in healthy females (fmean1=25.6; fmean2=25.9; fmax1=28.2; fmax2=28.7) and determined cut-off values for the diagnosis of ME/CFS in patients 20-39 years and 40-59 years respectively.^25^ Furthermore, we calculated the fatigue ratio as correlates of decreased force after repeated measurements and the recovery ratio indicating lower force during the second measurement.^25^ We decided to investigate multiple laboratory parameters, which have previously been associated with postinfectious fatigue syndromes including ferritin (reference range 13-150 μg/l), IL-8 in erythrocytes (reference <150 pg/ml), MBL (reference >50 ng/ml), ANAs (reference negative 1:0 dilution) and PO4 (reference range 0.87-1.45 mmol/l).^45^ They were determined at the Charité diagnostics laboratory (Labor Berlin GmbH, Berlin, Germany).

## Statistical analysis

Study data were collected and managed using REDCap, an electronic data collection system, and last accessed on December 13, 2022. Non-parametric rank-based ANOVA tests for factorial longitudinal data were used for data analysis. These tests are a generalization of the Wilcoxon-Mann-Whitney test for longitudinal data. We tested for group effects, time effects, and an interaction effect between these two effects. A group effect corresponds to a significant difference in the distribution of data between the PCS and the PCS-ME/CFS cohort, a time effect to a significant change in the data distribution over time, and an interaction effect to a temporal trend difference between the PCS and the PCS-ME/CFS cohort. Note that the reported effect sizes are relative (i.e., the probability of observing larger values for a given group at a given time). In a subsequent post-hoc analysis, Wilcoxon-Mann-Whitney tests were used to analyze group differences at individual time periods. The corresponding p-values were adjusted according to the Holm-Bonferroni correction. Correlation analyses were performed using Kendall’s Tau correlation. In general, p-values < 0.05 were considered statistically significant. Statistical testing was done in Prism version 9 and R version 4.2.1 with the packages tidyverse version 1.3 and nparLD version 2.2.

## Data Availability

All data produced in the present study are available upon reasonable request to the authors

## Acknowledgements

We thank Elena Steinle, Gritt Stoffels and Anja Hagemann for patient care and data management. We thank all patients who gave us their consent to publish their data in this study. We thank all members of the PaCOVID-19 collaborative study group.

## Funding

This work is supported by a grant from the Weidenhammer-Zoebele Foundation. The work of F. K. was supported by the Volkswagen Foundation.

## Data availability

Data is available upon reasonable request. Furthermore, the manuscript’s guarantor affirms that the paper is honest, accurate, and transparent; that no important aspects of the study have been omitted; and that any discrepancies from the study as planned have been explained. Dissemination to participants and related patient and public communities is encouraged by open access publication and citing the study on our site https://cfs.charite.de/. We are engaging with print and internet press, television, radio, news, and documentary program makers. All items of the STROBE checklist are covered in the paper.

## Author Contributions

C.S, C.K., and F.P. developed the concept of the study. L.M.-A. gave important input into study concept and objectives. A.F.K. and L.M.-A. were responsible for data curation and analyses of data. C.K., U.H., C.S., J.B.-S., L.M.-A., F.P. were involved in clinical investigation. A.F.L was involved in data transfer, data management and collection of further information for revision. H.F. was also involved in data transfer. C.K., J.B.-S. and C.S. validated the data. L.M. performed statistical analyses. A.F.L and L.M. were involved in data visualization. A.F.L and L.M.-A wrote the original draft of the paper. C.S. and J.B.-S. reviewed and edited the paper. All authors revised and approved the paper. The corresponding author attests that all listed authors meet authorship criteria and that no others meeting the criteria have been omitted. C.S. is the guarantor.

## Competing Interest Statements

The authors declare no competing interests.

## Notes

### Competing Interest Statement

The authors have declared no competing interest.

### Author Declarations

Ethics committee of Charite in accordance with the 1964 Declaration of Helsinki and its later amendments (EA2/006/20) gave ethical approval for this work

