## Supplementary Material for "Symptom persistence and biomarkers in post-COVID-19/chronic fatigue syndrome – results from a prospective observational cohort"

^1^ Charité – Universitätsmedizin Berlin, corporate member of Freie Universität Berlin, Humboldt Universität zu Berlin and Berlin Institute of Health, Max Delbrück for Molecular Medicine, Experimental and Clinical Research Center, 13125 Berlin, Germany

^2^ Charité – Universitätsmedizin Berlin, corporate member of Freie Universität Berlin, Humboldt Universität zu Berlin and Berlin Institute of Health, NeuroCure Research Center, 10117 Berlin, Germany

^3^ Charité – Universitätsmedizin Berlin, corporate member of Freie Universität Berlin, Humboldt Universität zu Berlin and Berlin Institute of Health, Department for Neurology with Experimental Neurology, 10117 Berlin, Germany

^4^ Charité - Universitätsmedizin Berlin, corporate member of Freie Universität Berlin and Humboldt-Universität zu Berlin and Institute of Biometry and Clinical Epidemiology, 10117 Berlin, Germany

^5^ Charité – Universitätsmedizin Berlin, corporate member of Freie Universität Berlin, Humboldt Universität zu Berlin and Berlin Institute of Health, Institute of Medical Immunology, 13353 Berlin, Germany

^6^Charité - Universitätsmedizin Berlin, corporate member of Freie Universität Berlin, Humboldt-Universität zu Berlin, and Berlin Institute of Health, Regenerative Immunology and Aging, BIH Center for Regenerative Therapies, 13353 Berlin, Germany

^7^Si-M / “Der Simulierte Mensch” a science framework of Technische Universität Berlin and Charité - Universitätsmedizin Berlin, 10117 Berlin, Germany

^+^ Equal contribution

^*^ Corresponding author

### Supplementary Tables

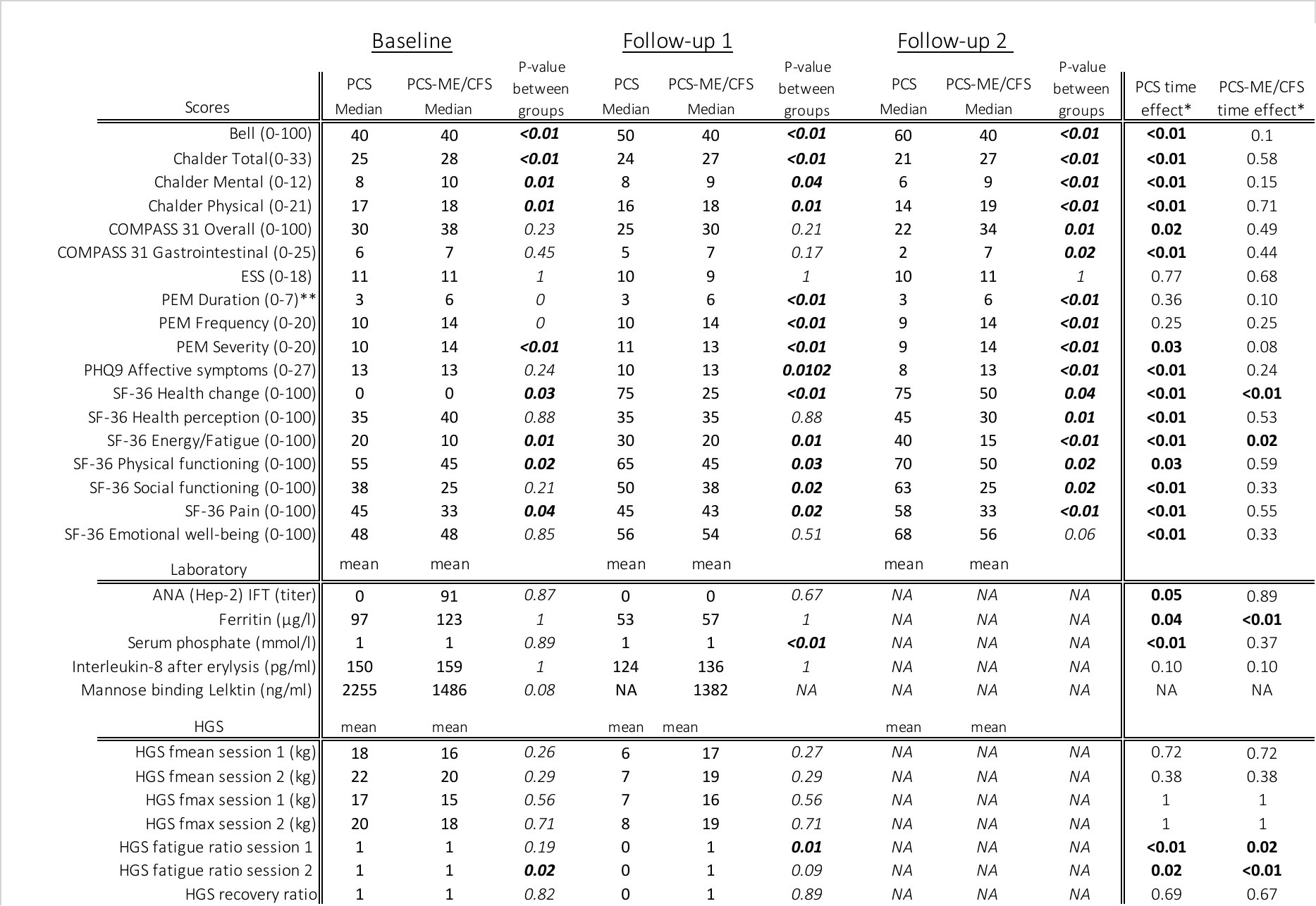
**Supplementary Table 1**

**Supp. Tab. 1:** Median values for symptom scores, mean values for biomarkers, and hand grip strength (HGS) for all three time periods. Group differences reported as adjusted p-values and significant results (< 0.05) are marked in bold. * Adjusted p-values for significant changes in data distribution over time. ** PEM Duration in total hours (0=0 hrs; 1=<1hr; 2=2-3hrs; 3=4-10hrs; 4=11-13hrs; 5=14-23hrs; 6=>24hrs; 7=>48hrs)

### Supplementary Figures

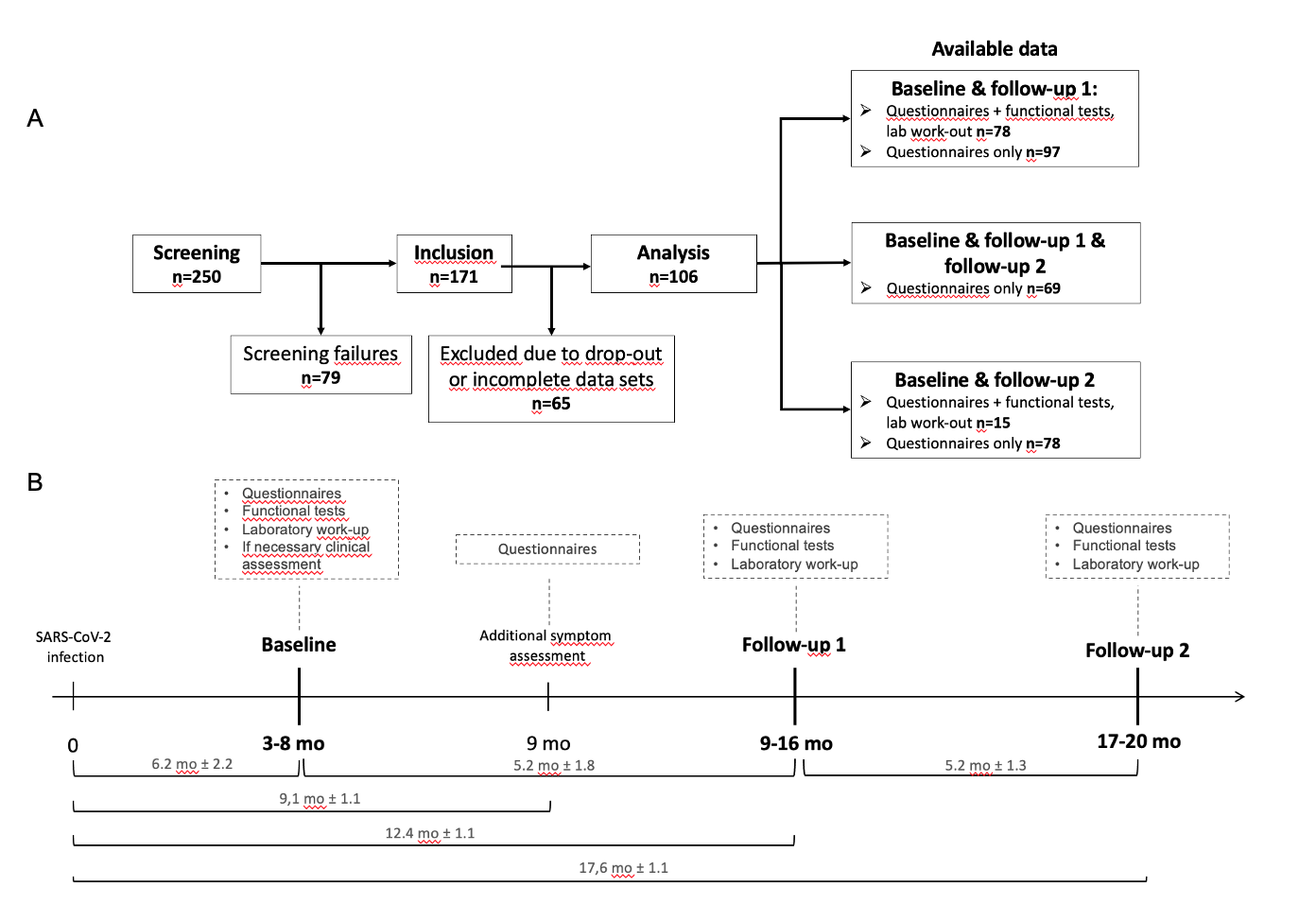
**Supplementary Fig. 1**

**Fig. 1: Participant flow chart and study design.** **a,** participant flow chart showing available data for screening and follow-up data collection, **b**, timeline depicting study visits including assessments conducted in relation to SARS-CoV-2 infection**.** Time intervals are reported as mean ± range; mo = months.

**Supplementary Fig. 2**

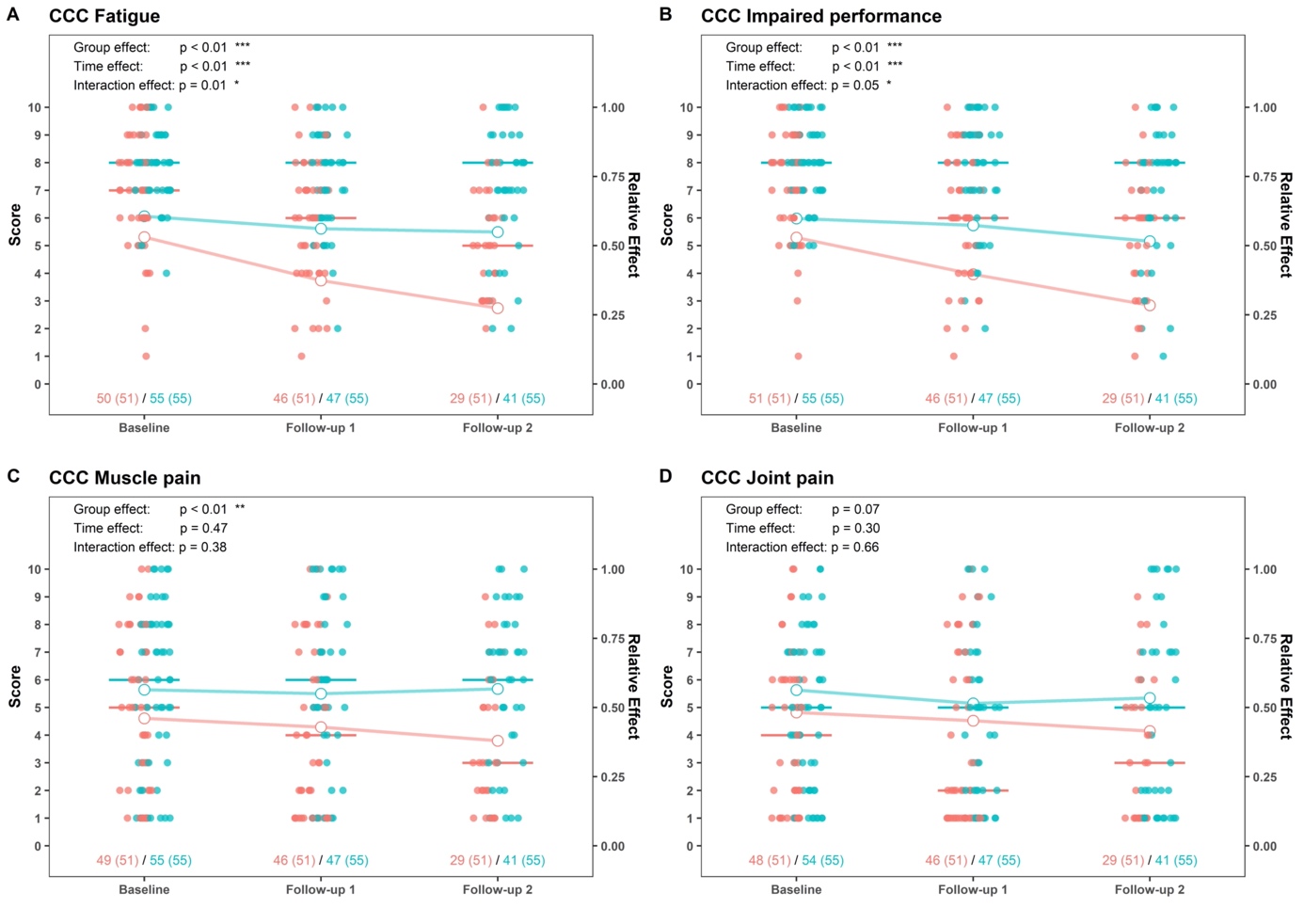

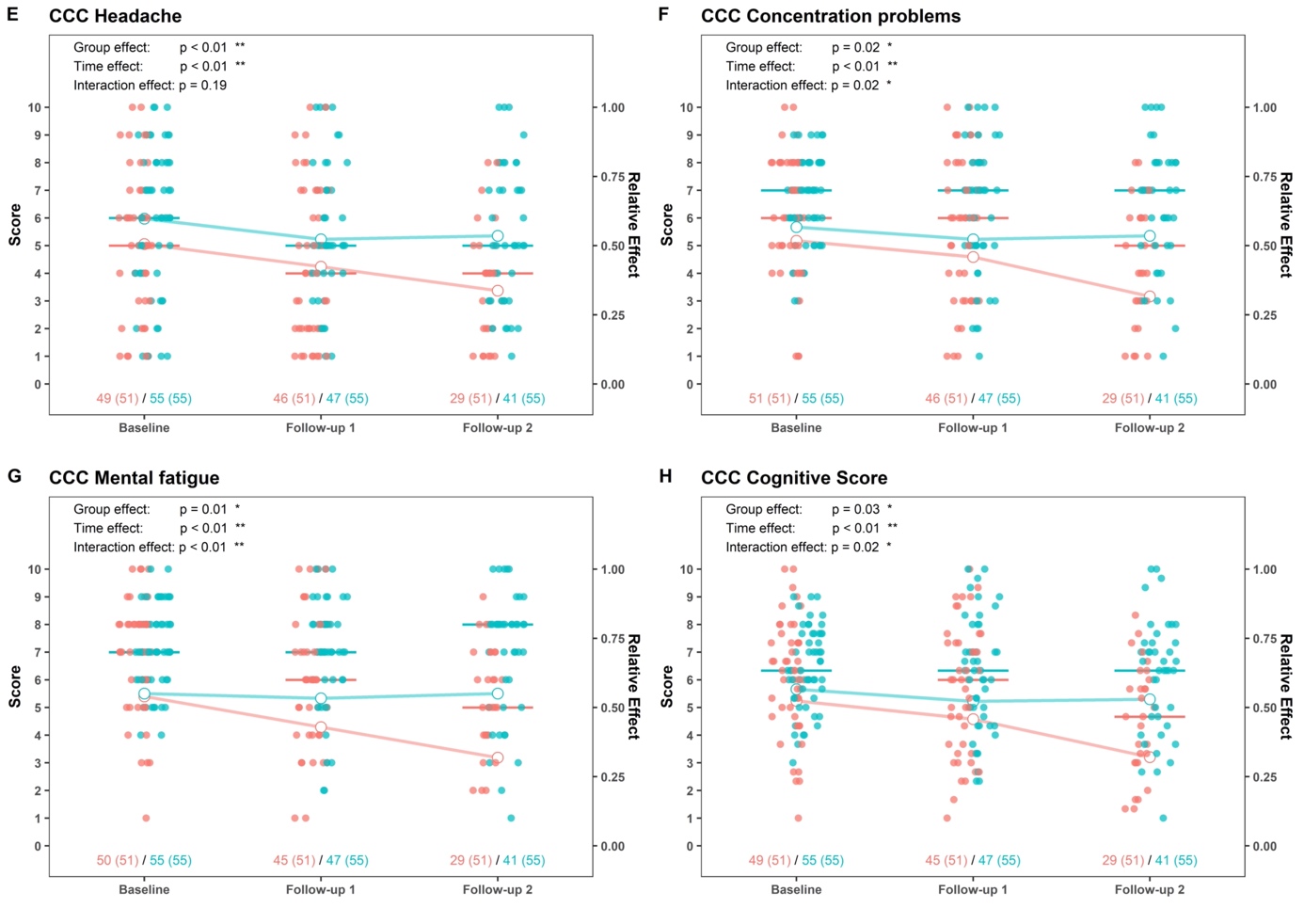

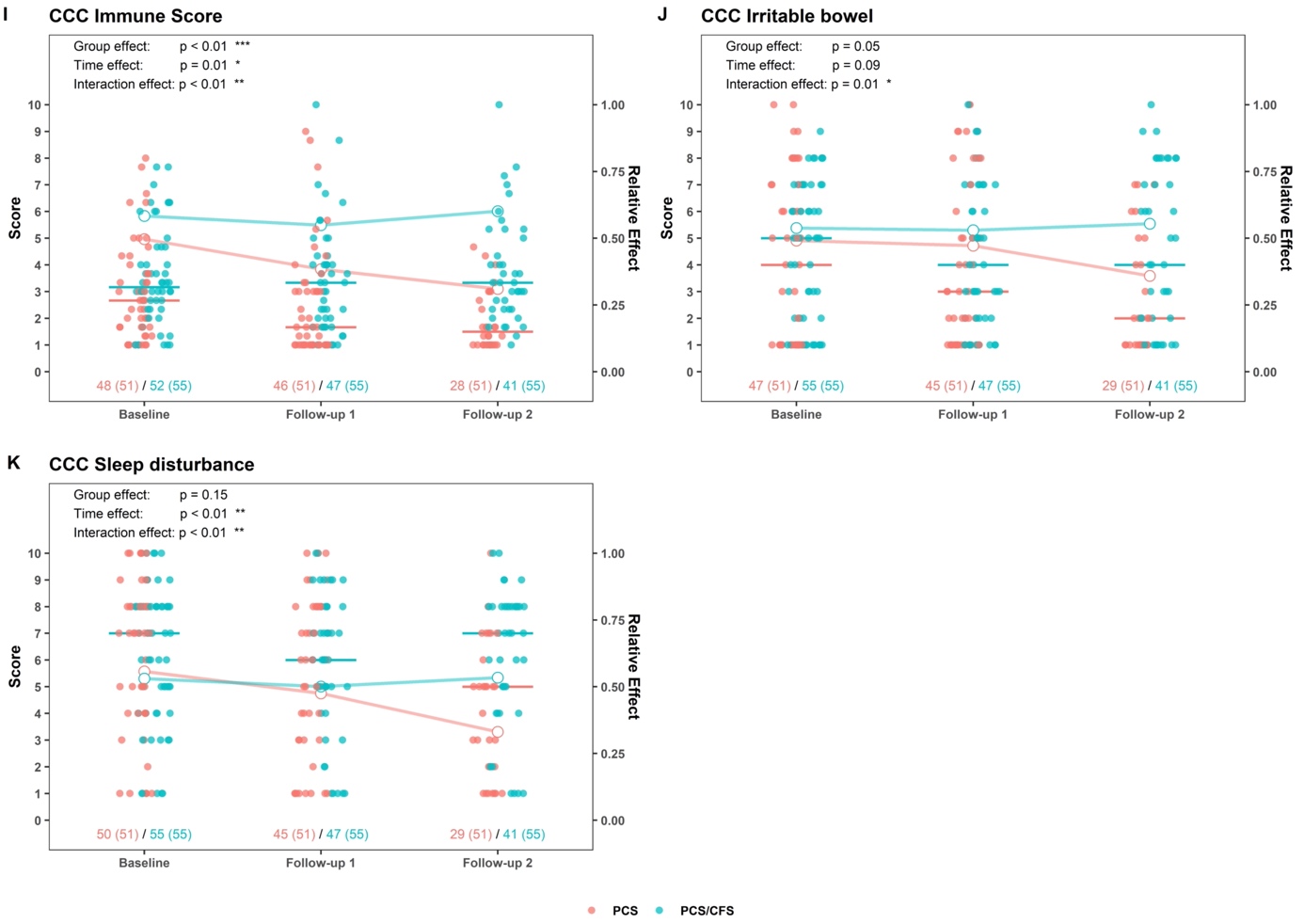

**Supp. Fig. 2: Symptom severity according to the quantified Canadian Consensus Criteria (CCCq).** **a,** fatigue, **b,** impaired performance, **c**, muscle pain, **d**, joint pain, **e**, headache, **f**, concentration problems, **g**, mental fatigue, **h**, cognitive score, **i**, immune score, **j**, irritable bowl, **k**, sleep disturbance. 0 points (no symptoms) - 10 points (high symptom severity). Dots represent absolute score values (red for PCS, blue for PCS-ME/CFS) as quantified on the left Y axis. Bars depict group medians. Lines (red for PCS, blue for PCS-ME/CFS) depict main relative time, group, and interaction effects as quantified on the right Y axis. p ≤ 0.05 = *, p ≤ 0.01=**, p ≤ 0.001=***, p ≤ 0.0001=****.
